# Protection offered by mRNA-1273 versus BNT162b2 vaccines against SARS-CoV-2 infection and severe COVID-19 in Qatar

**DOI:** 10.1101/2021.11.12.21266250

**Authors:** Laith J. Abu-Raddad, Hiam Chemaitelly, Houssein H. Ayoub, Patrick Tang, Mohammad R. Hasan, Peter Coyle, Hadi M. Yassine, Fatiha M. Benslimane, Hebah A. Al-Khatib, Zaina Al-Kanaani, Einas Al-Kuwari, Andrew Jeremijenko, Anvar Hassan Kaleeckal, Ali Nizar Latif, Riyazuddin Mohammad Shaik, Hanan F. Abdul-Rahim, Gheyath K. Nasrallah, Mohamed Ghaith Al-Kuwari, Adeel A. Butt, Hamad Eid Al-Romaihi, Mohamed H. Al-Thani, Abdullatif Al-Khal, Roberto Bertollini

## Abstract

**BACKGROUND:** Growing evidence suggests that COVID-19 vaccines differ in effectiveness against breakthrough infection or severe COVID-19, but vaccines have yet to be investigated in controlled studies that head-to-head compare immunity of one to another. This study compared protection offered by the mRNA-1273 (Moderna) vaccine with that of the BNT162b2 (Pfizer-BioNTech) vaccine in Qatar.

**METHODS:** In a population of 1,531,736 vaccinated persons, two matched retrospective cohort studies were designed and used to investigate differences in mRNA-1273 and BNT162b2 vaccine protection, after the first and second doses, from December 21, 2020 to October 20, 2021.

**RESULTS:** After dose 1, cumulative incidence of breakthrough infection was 0.79% (95% CI: 0.75-0.83%) for mRNA-1273-vaccinated individuals and 0.86% (95% CI: 0.82-0.90%) for BNT162b2-vaccinated individuals, 21 days post-injection. Adjusted hazard ratio (AHR) for breakthrough infection was 0.89 (95% CI: 0.83-0.95; p=0.001). AHR was constant in the first two weeks at 1, but it declined to 0.67 (95% CI: 0.57-0.77; p<0.001) in the third week after dose 1. AHR for any severe, critical, or fatal COVID-19 was 0.71 (95% CI: 0.53-0.95; p=0.020). After dose 2, cumulative incidence was 0.59% (95% CI: 0.55-0.64%) for mRNA-1273-vaccinated individuals and 0.84% (95% CI: 0.79-0.89%) for BNT162b2-vaccinated individuals, 180 days post-injection. AHR for breakthrough infection was 0.69 (95% CI: 0.63-0.75; p<0.001) and was largely constant over time after dose 2. AHR for any severe, critical, or fatal COVID-19 was 0.37 (95% CI: 0.10-1.41; p=0.147).

**CONCLUSIONS:** mRNA-1273 vaccination is associated with lower SARS-CoV-2 breakthrough infection and COVID-19 hospitalization and death than BNT162b2 vaccination, but the number of hospitalizations and deaths was exceedingly small for both vaccines. Both vaccines demonstrated strikingly similar patterns of build-up of protection after the first dose and waning of protection after the second dose.

## Introduction

Protection offered by Coronavirus Disease 2019 (COVID-19) vaccines against severe acute respiratory syndrome coronavirus 2 (SARS-CoV-2) infection and severe COVID-19 has been investigated in randomized clinical trials^1,2^ and in observational studies.^3^ A growing number of studies suggests that these vaccines differ in effectiveness in preventing breakthrough infection or severe COVID-19.^3-11^ However, protection conferred by these vaccines has not been investigated in controlled studies that directly compare one to another. In this study, we compared the protection conferred by the mRNA-1273^1^ (Moderna) vaccine to that of the BNT162b2^2^ (Pfizer-BioNTech) vaccine in Qatar, using a matched cohort study design that allows a head-to-head comparison of protection of these two mRNA vaccines.

## Methods

Hamad Medical Corporation and Weill Cornell Medicine-Qatar Institutional Review Boards approved this retrospective study with waiver of informed consent, since analysis was done on routinely collected data.

## Data sources, study population, and study design

This study analyzed the national, federated databases for SARS-CoV-2 infection in the resident population of Qatar, compiled at Hamad Medical Corporation, the main public healthcare provider and the nationally designated provider for all COVID-19 healthcare needs. These databases were constructed through a national electronic platform for health records designed to capture all SARS-CoV-2-related data. These include all polymerase chain reaction (PCR) testing, vaccination records, COVID-19 hospitalizations, infection severity and mortality classifications per World Health Organization (WHO) guidelines,^12,13^ in addition to sex, age, and nationality information retrieved from the national registry. Further descriptions of these national databases can be found in previous studies.^6,8,9,14-19^ Qatar has young, diverse demographics. Only 9% of the population are ≥50 years of age and 89% are resident expatriates from more than 150 countries.^14,20^

Qatar launched its mass COVID-19 immunization campaign on December 21, 2020, first using the BNT162b2^2^ vaccine,^6,8^ and three months later it added the mRNA-1273^1^ vaccine.^8^ Immunization with both vaccines followed the US Food and Drug Administration-approved dosing schedule.^1,2^ Vaccination was scaled up in phases to prioritize those most vulnerable to infection or to severe COVID-19.^19^ As of October 26, 2021, it is estimated that >80% of Qatar’s population had received both doses of either mRNA-1273 or BNT162b2.^21^ During a vaccination drive spanning nearly a year, Qatar experienced two epidemic waves predominated by the Alpha^22^ (B.1.1.7) and Beta^22^ (B.1.351) variants,^6,8,19,23-25^ but incidence since July 2021 has been predominated by the Delta^22^ (B.1.617.2) variant.^6,8,9,19,23-25^

Differences in incidence of breakthrough infection following administration of the first and second mRNA-1273 and BNT162b2 vaccine doses up to the study closing date (October 20, 2021) were investigated using two retrospective, matched cohort studies. The first study investigated incidence of breakthrough infection following administration of the first dose of each vaccine and before administration of the second dose. The second study investigated incidence of breakthrough infection in the months following administration of the second dose. Records of all mRNA-1273- and BNT162b2-vaccinated individuals in the Hamad Medical Corporation database, between December 21, 2020 and September 19, 2021 were reviewed. All persons with a record of SARS-CoV-2 infection prior to vaccination were excluded.

To control for differences in risk of exposure to the infection in Qatar^14,17,26-28^ and variant exposure,^6,8,9,19,23-25^ eligible mRNA-1273-vaccinated individuals were exact-matched in a 1:1 ratio by sex, 5-year age group, nationality, calendar week of the first dose in the first study, and calendar week of the second dose in the second study, to eligible BNT162b2-vaccinated individuals. Only matched samples were included in the analysis.

Follow-up in the first matched cohort study was from first dose vaccination up to the first occurrence of either PCR-confirmed infection, second dose vaccination, all-cause mortality, or end-of-study censoring. Follow-up in the second matched cohort study was from second dose vaccination up to the first occurrence of either PCR-confirmed infection, all-cause mortality, or end-of-study censoring.

Exposure in the first study was receiving only one dose of mRNA-1273 or BNT162b2 vaccines. Exposure in the second study was receiving the first and second doses of mRNA-1273 or BNT162b2 vaccines. All persons who received other vaccines or mixed different vaccines were excluded.

Investigated primary outcome in both studies was any PCR-positive nasopharyngeal swab regardless of the reason for PCR testing or presence of symptoms. Investigated secondary outcome in both studies was any severe,^12^ critical,^12^ or fatal^13^ breakthrough COVID-19 disease. Classification of COVID-19 case severity (acute-care hospitalizations),^12^ criticality (ICU hospitalizations),^12^ and fatality^13^ followed WHO guidelines, and assessments were made by trained medical personnel using individual chart reviews. Details of the COVID-19 severity, criticality, and fatality classification are found in Section 1 in Supplementary Appendix.

### Laboratory methods and classification of infections by variant type

Details of laboratory methods for real-time reverse-transcription PCR (RT-qPCR) testing are found in Section 2 in Supplementary Appendix. Methods for classification of infections by variant type using RT-qPCR variant screening^29^ of random positive clinical samples^23,25^ are included in Section 3 in Supplementary Appendix.

### Statistical analyses

Study populations were described using frequency distributions and measures of central tendency. Balance between study cohorts was assessed using standardized mean differences (SMDs), with SMD ≤0.1 indicating adequate matching.^30^

The Kaplan–Meier estimator method^31^ was used to estimate the cumulative incidence of infection. Cumulative incidence of infection was defined as the proportion of individuals at risk that were identified with a breakthrough infection during follow-up among all eligible individuals in each cohort. Equality of failure functions was assessed using the log-rank test. Standard errors of failure functions were used to derive 95% confidence intervals (CIs) of the absolute difference in cumulative incidence at different follow-up times.

Incidence rates and corresponding 95% CIs were estimated using a Poisson log-likelihood regression model with the STATA 17.0^32^ *stptime* command. Hazard ratios and corresponding 95% CIs were calculated using Cox regression adjusted for the matching factors with the STATA 17.0^32^ *stcox* command. Shoenfeld residuals and log-log plots confirmed adequacy of the proportional-hazards assumption. 95% CIs were not adjusted for multiplicity. Interactions were not considered. Subgroup analyses were performed to estimate adjusted hazard ratios at different follow-up times.

In all analyses, two-sided p<0.05 indicated statistical significance. Statistical analyses were conducted in STATA/SE version 17.0.^32^ STROBE checklist is in Table S1.

## Results

### Study population

Of 1,531,736 vaccinated persons, 491,875 mRNA-1273-vaccinated individuals and 845,326 BNT162b2-vaccinated individuals met the eligibility criteria for the study, assessing incidence of breakthrough infection after dose 1 (Figure 1). Also, of the 1,531,736 vaccinated persons, 443,416 mRNA-1273-vaccinated individuals and 799,836 BNT162b2-vaccinated individuals met the eligibility criteria for the study, assessing incidence of breakthrough infection after dose 2 (Figure 1).

**Figure 1.**
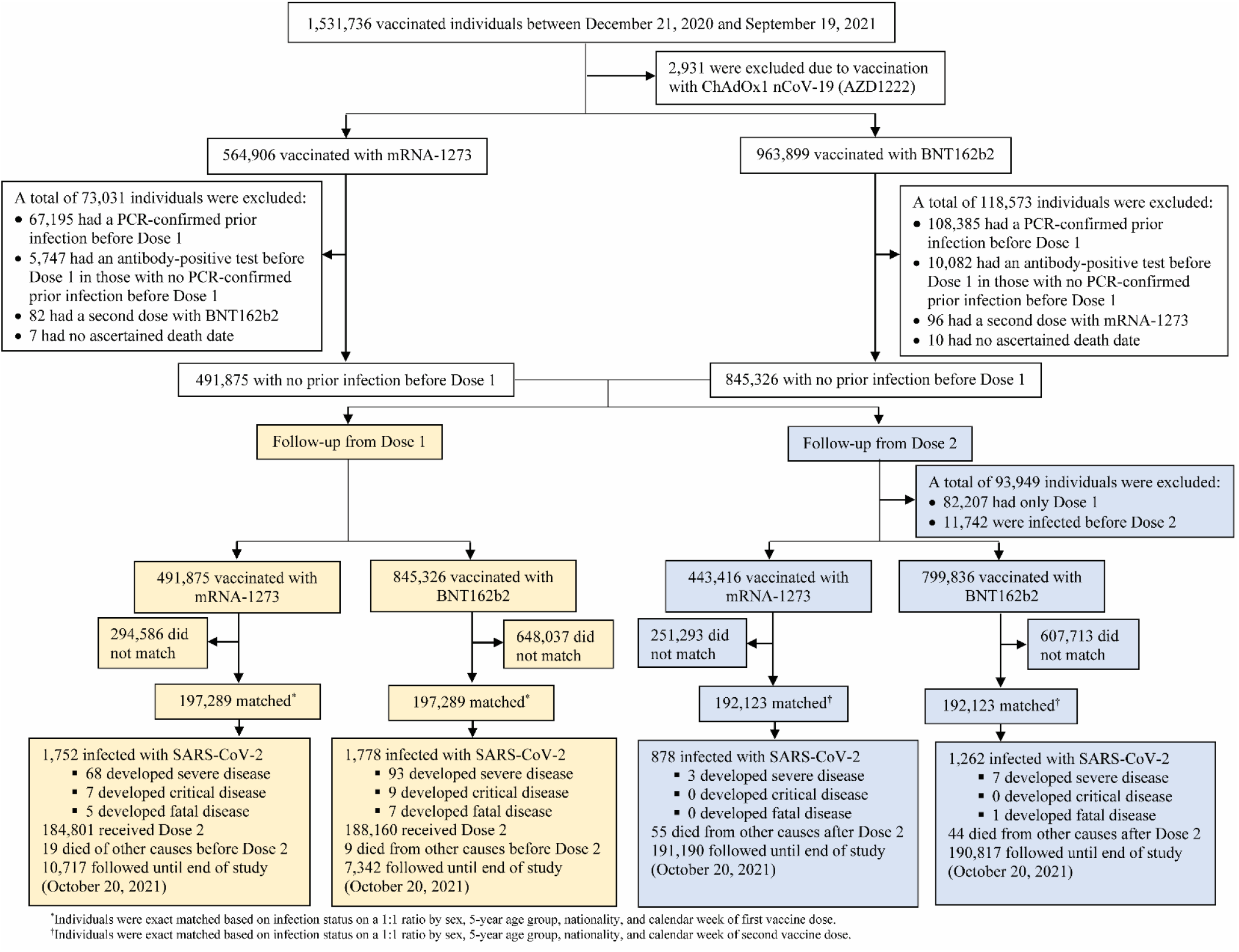
Development of cohorts in a study of SARS-CoV-2 breakthrough infections after vaccination with the mRNA-1273 and BNT162b2 vaccines.

Baseline characteristics of mRNA-1273- and BNT162b2-vaccinated cohorts that were followed up from the date of dose 1 are presented in Table 1. Prior to matching, there were small differences in sex, but large differences in age distribution, nationality, and calendar month of dose 1. These differences have arisen because the mass mRNA-1273 immunization campaign started three months after the BNT162b2 campaign; vaccination was prioritized to frontline health workers, persons with severe or multiple chronic conditions, specific occupational groups, such as teachers, and by age;^19^ all in a context in which there are associations between age, sex, nationality, and occupation.^14,17,26-28^

**Table 1.**
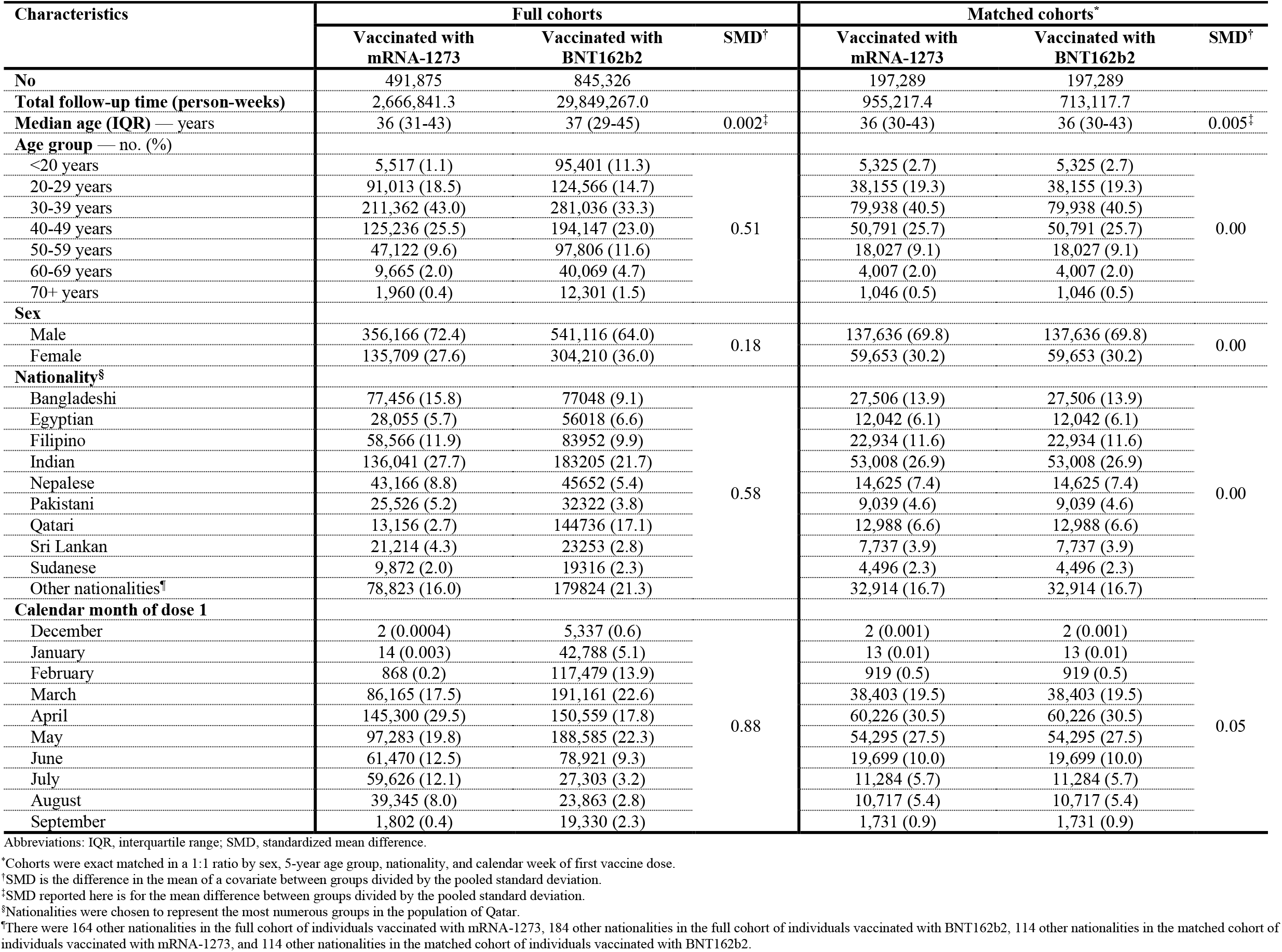
Baseline characteristics of cohorts that received the mRNA-1273 and BNT162b2 vaccines and were followed from the date of the first vaccine dose.

Baseline characteristics of the mRNA-1273- and BNT162b2-vaccinated cohorts that were followed up from the date of dose 2 are presented in Table 2. Similar differences were observed for the same reasons as for those followed up from the date of dose 1.

**Table 2.** Baseline characteristics of cohorts that received the mRNA-1273 and BNT162b2 vaccines and were followed from the date of the second vaccine dose.

| Characteristics | Full cohorts |  |  | Matched cohorts* |  |  |
| --- | --- | --- | --- | --- | --- | --- |
|  | Vaccinated with mRNA-1273 | Vaccinated with BNT162b2 | SMD† | Vaccinated with mRNA-1273 | Vaccinated with BNT162b2 | SMD† |
| **No** | 443,416 | 799,836 |  | 192,123 | 192,123 |  |
| **Total follow-up time (person-weeks)** | 8,247,683.6 | 18,910,917.6 |  | 3,894,874.6 | 3,886,201.2 |  |
| **Median age (IQR) — years** | 36 (31-44) | 37 (29-46) | 0.004‡ | 37 (31-44) | 37 (31-44) | 0.006‡ |
| **Age group — no. (%)** |  |  |  |  |  |  |
| <20 years | 5,260 (1.2) | 87,449 (10.9) | 0.49 | 4,869 (2.5) | 4,869 (2.5) | 0.00 |
| 20-29 years | 84,462 (19.1) | 116,312 (14.5) |  | 36,273 (18.9) | 36,273 (18.9) |  |
| 30-39 years | 179,186 (40.4) | 265,626 (33.2) |  | 71,959 (37.5) | 71,959 (37.5) |  |
| 40-49 years | 118,523 (26.7) | 184,969 (23.1) |  | 57,777 (30.1) | 57,777 (30.1) |  |
| 50-59 years | 44,847 (10.1) | 94,448 (11.8) |  | 16,945 (8.8) | 16,945 (8.8) |  |
| 60-69 years | 9,266 (2.1) | 39,029 (4.9) |  | 3,378 (1.8) | 3,378 (1.8) |  |
| 70+ years | 1,872 (0.4) | 12,003 (1.5) |  | 922 (0.5) | 922 (0.5) |  |
| **Sex** |  |  |  |  |  |  |
| Male | 314,035 (70.8) | 511,821 (64.0) | 0.15 | 131,959 (68.7) | 131,959 (68.7) | 0.00 |
| Female | 129,381 (29.2) | 288,015 (36.0) |  | 60,164 (31.3) | 60,164 (31.3) |  |
| **Nationality§** |  |  |  |  |  |  |
| Bangladeshi | 64,432 (14.5) | 71,384 (8.9) | 0.56 | 24,485 (12.7) | 24,485 (12.7) | 0.00 |
| Egyptian | 26,514 (6.0) | 52,406 (6.6) |  | 11,191 (5.8) | 11,191 (5.8) |  |
| Filipino | 55,403 (12.5) | 79,725 (10.0) |  | 24,712 (12.9) | 24,712 (12.9) |  |
| Indian | 124,550 (28.1) | 173,921 (21.7) |  | 53,260 (27.7) | 53,260 (27.7) |  |
| Nepalese | 35,320 (8.0) | 42,712 (5.3) |  | 14,383 (7.5) | 14,383 (7.5) |  |
| Pakistani | 22,441 (5.1) | 29,658 (3.7) |  | 8,399 (4.4) | 8,399 (4.4) |  |
| Qatari | 12,678 (2.9) | 139,350 (17.4) |  | 12,136 (6.3) | 12,136 (6.3) |  |
| Sri Lankan | 19,360 (4.4) | 21,920 (2.7) |  | 7,816 (4.1) | 7,816 (4.1) |  |
| Sudanese | 9,170 (2.1) | 18,191 (2.3) |  | 4,331 (2.3) | 4,331 (2.3) |  |
| Other nationalities¶ | 73,548 (16.6) | 170,569 (21.3) |  | 31,410 (16.4) | 31,410 (16.4) |  |
| **Calendar month of dose 2** |  |  |  |  |  |  |
| January | 1 (0.0002) | 13,722 (1.7) | 1.01 | 1 (0.0005) | 1 (0.0005) | 0.08 |
| February | 9 (0.002) | 46,671 (5.8) |  | 9 (0.005) | 9 (0.005) |  |
| March | 1,537 (0.4) | 182,292 (22.8) |  | 1,511 (0.8) | 1,614 (0.8) |  |
| April | 90,741 (20.5) | 121,098 (15.1) |  | 37,461 (19.5) | 31,937 (16.6) |  |
| May | 134,849 (30.4) | 196,884 (24.6) |  | 80,339 (41.8) | 85,760 (44.6) |  |
| June | 74,642 (16.8) | 130,170 (16.3) |  | 33,268 (17.3) | 34,040 (17.7) |  |
| July | 51,091 (11.5) | 71,311 (8.9) |  | 24,267 (12.6) | 24,195 (12.6) |  |
| August | 65,435 (14.8) | 24,126 (3.0) |  | 9,058 (4.7) | 8,274 (4.3) |  |
| September | 25,111 (5.7) | 13,562 (1.7) |  | 6,209 (3.2) | 6,293 (3.3) |  |

Abbreviations: IQR, interquartile range; SMD, standardized mean difference.
\*Cohorts were exact matched in a 1:1 ratio by sex, 5-year age group, nationality, and calendar week of second vaccine dose.
<sup>1</sup>SMD is the difference in the mean of a covariate between groups divided by the pooled standard deviation.
<sup>2</sup>SMD reported here is for the mean difference between groups divided by the pooled standard deviation.
<sup>3</sup>Nationalities were chosen to represent the most numerous groups in the population of Qatar.
<sup>4</sup>There were 164 other nationalities in the full cohort of individuals vaccinated with mRNA-1273, 184 other nationalities in the full cohort of individuals vaccinated with BNT162b2, 115 other nationalities in the matched cohort of individuals vaccinated with mRNA-1273, and 115 other nationalities in the matched cohort of individuals vaccinated with BNT162b2.

Median time elapsed between the first and second vaccine dose was 28 days (interquartile range (IQR): 28-31 days) for mRNA-1273 and 21 days (IQR: 21-22 days) for BNT162b2. Of mRNA-1273- and BNT162b2-vaccinated individuals, 74.9% and 97.4%, respectively, received the second dose ≤30 days after the first dose.

### Breakthrough infection incidence after dose 1

Of 491,875 mRNA-1273-vaccinated individuals with no record of prior infection before dose 1, 197,289 were exact-matched to 197,289 BNT162b2-vaccinated individuals with no record of prior infection (Figure 1). These matched cohorts were balanced on the matching factors (Table 1). The median date of dose 1 was April 28, 2021. Among mRNA-1273- and BNT162b2-vaccinated individuals, 7.4% and 6.5% had a PCR test done during follow-up, respectively. The higher proportion among those vaccinated with mRNA-1273 is a consequence of longer follow-up with the one-week delayed dose 2 schedule, compared to those BNT162b2 vaccinated.

Among mRNA-1273-vaccinated individuals, 1,752 breakthrough infections were recorded after dose 1 and before dose 2 at a median follow-up of 9 days (IQR: 6-14 days; Figure 1; Table 3). Of these infections, 68 progressed to severe COVID-19 disease, 7 to critical disease, and 5 to COVID-19 death.

**Table 3.** Baseline characteristics of breakthrough-infected individuals after the first vaccine dose and after the second vaccine dose in the matched cohorts of those vaccinated with mRNA-1273 and BNT162b2.

| Characteristics | Infected after Dose 1 |  | SMD<sup>‡</sup> | Infected after Dose 2 |  | SMD<sup>‡</sup> |
| --- | --- | --- | --- | --- | --- | --- |
|  | Vaccinated with mRNA-1273<sup>*</sup> | Vaccinated with BNT162b2<sup>*</sup> |  | Vaccinated with mRNA-1273<sup>†</sup> | Vaccinated with BNT162b2<sup>†</sup> |  |
| No | 1,752 | 1,778 |  | 878 | 1,262 |  |
| Median time from start of follow-up (dose) to infection (IQR) — days | 9 (6-14) | 10 (6-15) |  | 89 (29-123) | 86 (23-122) |  |
| Median age (IQR) — years | 40 (32-38) | 40 (32-38) | 0.002<sup>§</sup> | 36 (30-42) | 37 (30-44) | 0.05<sup>§</sup> |
| Age group — no. (%) |  |  |  |  |  |  |
| <20 years | 25 (1.4) | 37 (2.1) | 0.08 | 20 (2.3) | 33 (2.6) | 0.12 |
| 20-29 years | 264 (15.1) | 240 (13.5) |  | 175 (19.9) | 254 (20.1) |  |
| 30-39 years | 566 (32.3) | 608 (34.2) |  | 355 (40.4) | 470 (37.2) |  |
| 40-49 years | 526 (30.0) | 518 (29.1) |  | 243 (27.7) | 348 (27.6) |  |
| 50-59 years | 276 (15.8) | 270 (15.2) |  | 62 (7.1) | 126 (10.0) |  |
| 60-69 years | 72 (4.1) | 84 (4.7) |  | 20 (2.3) | 27 (2.1) |  |
| 70+ years | 23 (1.3) | 21 (1.2) |  | 3 (0.3) | 4 (0.3) |  |
| Sex |  |  |  |  |  |  |
| Male | 1,147 (65.5) | 1,108 (62.3) | 0.07 | 496 (56.5) | 748 (59.3) | 0.06 |
| Female | 605 (34.5) | 670 (37.7) |  | 382 (43.5) | 514 (40.7) |  |
| Nationality<sup>†</sup> |  |  |  |  |  |  |
| Bangladeshi | 150 (8.6) | 108 (6.1) | 0.13 | 82 (9.3) | 100 (7.9) | 0.11 |
| Egyptian | 131 (7.5) | 162 (9.1) |  | 81 (9.2) | 107 (8.5) |  |
| Filipino | 234 (13.4) | 250 (14.1) |  | 85 (9.7) | 125 (9.9) |  |
| Indian | 427 (24.4) | 467 (26.3) |  | 219 (24.9) | 289 (22.9) |  |
| Nepalese | 83 (4.7) | 76 (4.3) |  | 24 (2.7) | 40 (3.2) |  |
| Pakistani | 60 (3.4) | 56 (3.2) |  | 38 (4.3) | 52 (4.1) |  |
| Qatari | 230 (13.1) | 246 (13.8) |  | 140 (16.0) | 241 (19.1) |  |
| Sri Lankan | 61 (3.5) | 62 (3.5) |  | 22 (2.5) | 27 (2.1) |  |
| Sudanese | 54 (3.1) | 50 (2.8) |  | 17 (1.9) | 27 (2.1) |  |
| Other nationalities<sup>**</sup> | 322 (18.4) | 301 (16.9) |  | 170 (19.4) | 254 (20.1) |  |
| Calendar month of vaccine dose<sup>††</sup> |  |  |  |  |  |  |
| January | 0 (0) | 1 (0.06) | 0.12 | -- | -- | 0.10 |
| February | 5 (0.3) | 15 (0.8) |  | -- | -- |  |
| March | 959 (54.7) | 997 (56.1) |  | 31 (3.5) | 51 (4.0) |  |
| April | 675 (38.5) | 631 (35.5) |  | 359 (40.9) | 512 (40.6) |  |
| May | 79 (4.5) | 91 (5.1) |  | 365 (41.6) | 530 (42.0) |  |
| June | 16 (0.9) | 18 (1.0) |  | 75 (8.5) | 110 (8.7) |  |
| July | 11 (0.6) | 11 (0.6) |  | 37 (4.2) | 53 (4.2) |  |
| August | 6 (0.3) | 14 (0.8) |  | 6 (0.7) | 5 (0.4) |  |
| September | 1 (0.06) | 0 (0) |  | 5 (0.6) | 1 (0.08) |  |

Abbreviations: IQR, interquartile range; SMD, standardized mean difference.
<sup>\*</sup>Cohorts were exact matched in a 1:1 ratio by sex, 5-year age group, nationality, and calendar week of first vaccine dose.
<sup>†</sup>Cohorts were exact matched in a 1:1 ratio by sex, 5-year age group, nationality, and calendar week of second vaccine dose.
<sup>‡</sup>SMD is the difference in the mean of a covariate between groups divided by the pooled standard deviation.
<sup>§</sup>SMD reported here is for the mean difference between groups divided by the pooled standard deviation.
<sup>\*\*</sup>Nationalities were chosen to represent the most numerous groups in the population of Qatar.
<sup>\*\*\*</sup>There were 34 other nationalities in mRNA-1273-vaccinated individuals with breakthrough infection after dose 1, 51 other nationalities in BNT162b2-vaccinated individual with breakthrough infection after dose 1, 28 other nationalities in mRNA-1273-vaccinated individuals with breakthrough infection after dose 2, and 41 other nationalities in BNT162b2-vaccinated individuals with breakthrough infection after dose 2.
<sup>††</sup>Calendar month of vaccine dose was for dose 1 in the study assessing breakthrough infections after dose 1, and for dose 2 in the study assessing breakthrough infections after dose 2.

Among BNT162b2-vaccinated individuals, 1,778 breakthrough infections were recorded after dose 1 and before dose 2 at a median follow-up of 10 days (IQR: 6-15 days; Figure 1; Table 3). Of these infections, 93 progressed to severe COVID-19 disease, 9 to critical disease, and 7 to COVID-19 death.

There were minimal differences in median age, age distribution, sex, nationality, and calendar month of dose 1 between mRNA-1273 and BNT162b2 breakthrough infections (Table 3). Most breakthrough infections in both vaccinated cohorts occurred during the large Beta variant wave between March and May 2021.^6,8,9,19,23-25^ The median date of breakthrough infection was April 14, 2021. Only a minority of infections occurred after June 2021, when the Delta variant predominated.^6,8,9,19,23-25^

Cumulative incidence of breakthrough infection after dose 1 was estimated at 0.79% (95% CI: 0.75-0.83%) for mRNA-1273-vaccinated individuals and at 0.86% (95% CI: 0.82-0.90%) for BNT162b2-vaccinated individuals, at 21 days of follow-up (Figure 2A). Cumulative incidence curves of the two vaccinated cohorts started to diverge at the beginning of the third week after dose 1. However, both curves followed a similar pattern in which cumulative incidence started to plateau by the end of the third week of follow-up, consistent with attainment of considerable protection against infection.

**Figure 2.**
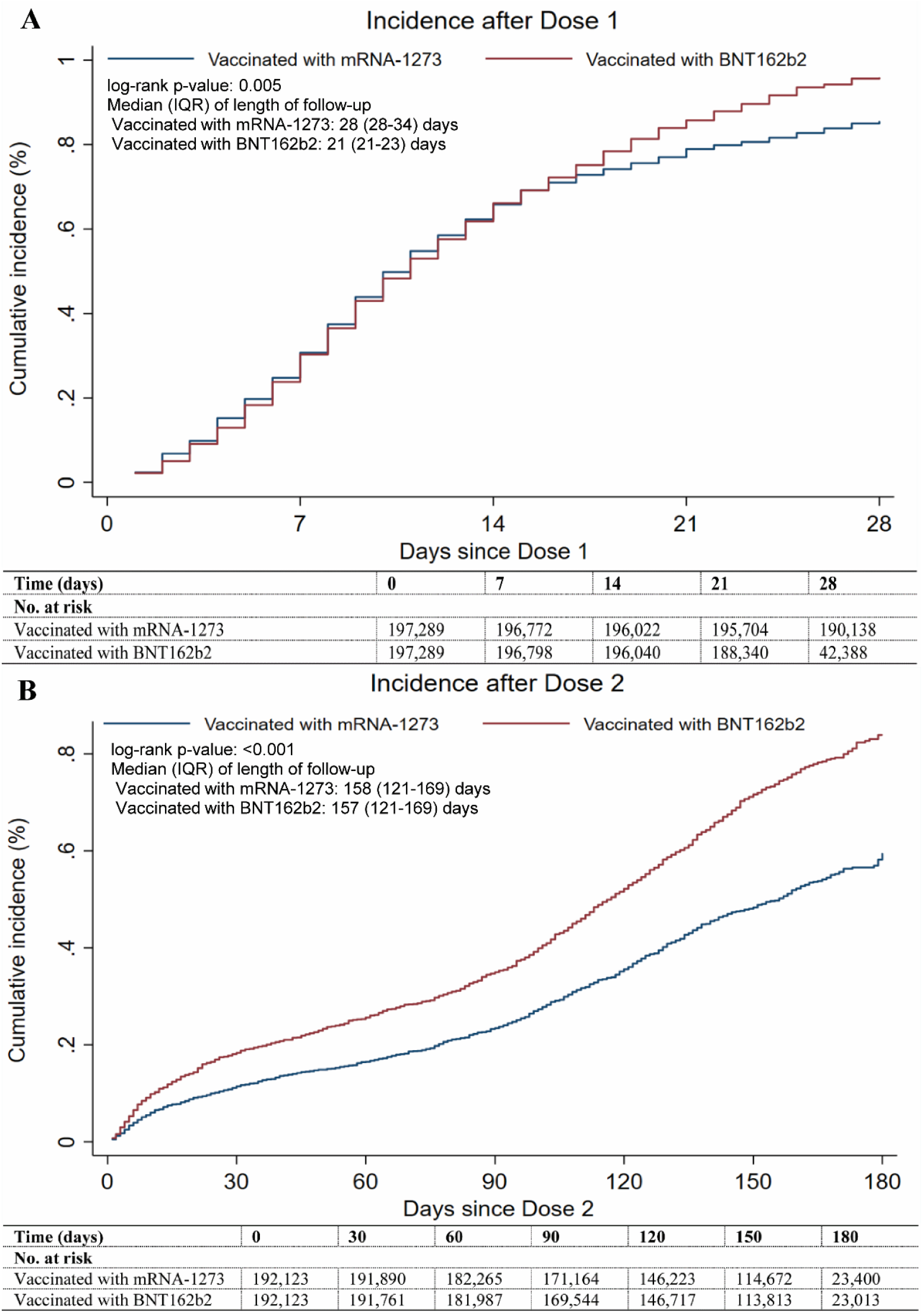
Cumulative incidence of breakthrough infection among matched cohorts of individuals vaccinated with mRNA-1273 and BNT162b2.

Breakthrough infection incidence rates among mRNA-1273- and BNT162b2-vaccinated individuals were estimated at 18.34 (95% CI: 17.50-19.22) and 24.93 (95% CI: 23.80-26.12) per 10,000 person-weeks, respectively. The overall hazard ratio for breakthrough infection after dose 1, adjusted for 5-year age group, sex, nationality group, and calendar week of dose 1, was estimated at 0.89 (95% CI: 0.83-0.95; p=0.001). The adjusted hazard ratio was stable in the first and second weeks after dose 1, at 1.04 (95% CI: 0.92-1.18; p=0.538) and 0.99 (95% CI: 0.89-1.09; p=0.801), respectively, but it declined to 0.67 (95% CI: 0.57-0.77; p<0.001) in the third week (Table 4). The overall adjusted hazard ratio for any severe, critical, or fatal COVID-19 disease after dose 1 was estimated at 0.71 (95% CI: 0.53-0.95; p=0.020).

**Table 4.**
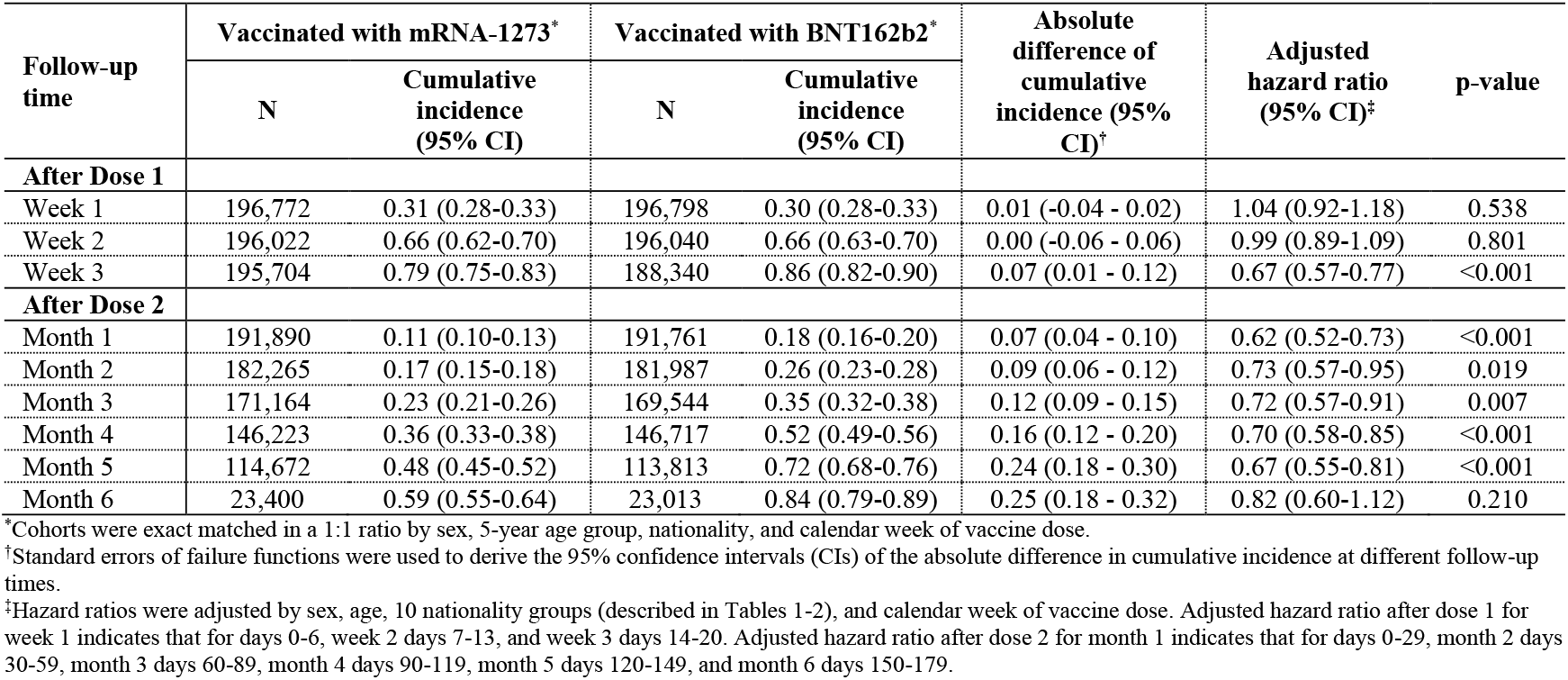
Adjusted hazard ratio for breakthrough infection by week of follow-up after the first vaccine dose, and by month of follow-up after the second vaccine dose, comparing matched cohorts of individuals vaccinated with mRNA-1273 and BNT162b2.

| Follow-up time | Vaccinated with mRNA-1273* |  | Vaccinated with BNT162b2* |  | Absolute difference of cumulative incidence (95% CI)<sup>†</sup> | Adjusted hazard ratio (95% CI)<sup>‡</sup> | p-value |
| --- | --- | --- | --- | --- | --- | --- | --- |
|  | N | Cumulative incidence (95% CI) | N | Cumulative incidence (95% CI) |  |  |  |
| **After Dose 1** |  |  |  |  |  |  |  |
| Week 1 | 196,772 | 0.31 (0.28-0.33) | 196,798 | 0.30 (0.28-0.33) | 0.01 (-0.04 - 0.02) | 1.04 (0.92-1.18) | 0.538 |
| Week 2 | 196,022 | 0.66 (0.62-0.70) | 196,040 | 0.66 (0.63-0.70) | 0.00 (-0.06 - 0.06) | 0.99 (0.89-1.09) | 0.801 |
| Week 3 | 195,704 | 0.79 (0.75-0.83) | 188,340 | 0.86 (0.82-0.90) | 0.07 (0.01 - 0.12) | 0.67 (0.57-0.77) | <0.001 |
| **After Dose 2** |  |  |  |  |  |  |  |
| Month 1 | 191,890 | 0.11 (0.10-0.13) | 191,761 | 0.18 (0.16-0.20) | 0.07 (0.04 - 0.10) | 0.62 (0.52-0.73) | <0.001 |
| Month 2 | 182,265 | 0.17 (0.15-0.18) | 181,987 | 0.26 (0.23-0.28) | 0.09 (0.06 - 0.12) | 0.73 (0.57-0.95) | 0.019 |
| Month 3 | 171,164 | 0.23 (0.21-0.26) | 169,544 | 0.35 (0.32-0.38) | 0.12 (0.09 - 0.15) | 0.72 (0.57-0.91) | 0.007 |
| Month 4 | 146,223 | 0.36 (0.33-0.38) | 146,717 | 0.52 (0.49-0.56) | 0.16 (0.12 - 0.20) | 0.70 (0.58-0.85) | <0.001 |
| Month 5 | 114,672 | 0.48 (0.45-0.52) | 113,813 | 0.72 (0.68-0.76) | 0.24 (0.18 - 0.30) | 0.67 (0.55-0.81) | <0.001 |
| Month 6 | 23,400 | 0.59 (0.55-0.64) | 23,013 | 0.84 (0.79-0.89) | 0.25 (0.18 - 0.32) | 0.82 (0.60-1.12) | 0.210 |

\*Cohorts were exact matched in a 1:1 ratio by sex, 5-year age group, nationality, and calendar week of vaccine dose.
<sup>†</sup>Standard errors of failure functions were used to derive the 95% confidence intervals (CIs) of the absolute difference in cumulative incidence at different follow-up times.
<sup>‡</sup>Hazard ratios were adjusted by sex, age, 10 nationality groups (described in Tables 1-2), and calendar week of vaccine dose. Adjusted hazard ratio after dose 1 for week 1 indicates that for days 0-6, week 2 days 7-13, and week 3 days 14-20. Adjusted hazard ratio after dose 2 for month 1 indicates that for days 0-29, month 2 days 30-59, month 3 days 60-89, month 4 days 90-119, month 5 days 120-149, and month 6 days 150-179.

### Breakthrough infection incidence after dose 2

Of 443,416 mRNA-1273-vaccinated individuals with no record of prior infection before dose 2, 192,123 were exact-matched to 192,123 BNT162b2-vaccinated individuals with no record of prior infection (Figure 1). These matched cohorts were balanced on the matching factors (Table 2). The median date of dose 2 was May 15, 2021. Among mRNA-1273- and BNT162b2-vaccinated individuals, 44.4% and 43.0% had a PCR test done during follow-up, respectively.

Among mRNA-1273-vaccinated individuals, 878 breakthrough infections were recorded after dose 2 at a median follow-up of 89 days (IQR: 29-123 days; Figure 1; Table 3). Of these infections, 3 progressed to severe COVID-19 disease and none to critical or fatal disease. Among BNT162b2-vaccinated individuals, 1,262 breakthrough infections were recorded after dose 2 at a median follow-up of 86 days (IQR: 23-122 days; Figure 1; Table 3). Of these infections, 7 progressed to severe COVID-19 disease, none to critical disease, and 1 to COVID-19 death.

There were minimal differences in median age, age distribution, sex, nationality, and calendar month of dose 2 between mRNA-1273 and BNT162b2 breakthrough infections (Table 3). Most breakthrough infections in both vaccinated cohorts occurred during the Delta-dominated low-incidence phase after June 2021.^6,8,9,19,23-25^ The median date of breakthrough infection was July 25, 2021. For both vaccinated cohorts, breakthrough infections tended to occur among those who had received their vaccination earlier (Figure 2B).

Cumulative incidence of breakthrough infection after dose 2 was estimated at 0.59% (95% CI: 0.55-0.64%) for mRNA-1273-vaccinated individuals and at 0.84% (95% CI: 0.79-0.89%) for BNT162b2-vaccinated individuals, at 180 days of follow-up (Figure 2B). Cumulative incidence curves of the two vaccinated cohorts started to diverge immediately after dose 2, but both curves followed a similar pattern. Cumulative incidence started to plateau around 10 days after dose 2, consistent with attainment of the highest protection against infection after two vaccine doses. However, at around 90 days after dose 2, both curves started to bend upward, at a time of low infection incidence in the wider population, consistent with progressive waning of vaccine protection.

Breakthrough infection incidence rates among mRNA-1273- and BNT162b2-vaccinated individuals were estimated at 2.25 (95% CI: 2.11-2.41) and 3.25 (95% CI: 3.07-3.43) per 10,000 person-weeks, respectively. The overall hazard ratio for breakthrough infection after dose 2, adjusted for 5-year age group, sex, nationality group, and calendar week of dose 2, was estimated at 0.69 (95% CI: 0.63-0.75; p<0.001). The adjusted hazard ratio was largely stable over time after dose 2 at about this value (Table 4). The overall adjusted hazard ratio for any severe, critical, or fatal COVID-19 disease after dose 2 was estimated at 0.37 (95% CI: 0.10-1.41; p=0.147).

## Discussion

Significant differences were observed in protection offered by the mRNA-1273 vaccine versus that of the BNT162b2 vaccine, against both breakthrough infection and severe COVID-19. These differences started to emerge in the third week after the first dose. Incidence of breakthrough infection in the third week after the first dose, and among persons who received both doses, was 30% lower among mRNA-1273-vaccinated persons compared to that among BNT162b2-vaccinated persons. After the first dose, incidence of severe, critical, or fatal COVID-19 cases was 30% lower among mRNA-1273-vaccined persons than among BNT162b2-vaccinated persons. After the second dose, incidence of these cases was also lower among mRNA-1273-vaccined persons, but the difference did not reach statistical significance, perhaps because of the small number of cases. The number of severe, critical, or fatal COVID-19 cases was exceedingly small among persons fully vaccinated with either vaccine, indicating robust protection against severe forms of COVID-19.

Despite these differences, both vaccines demonstrated strikingly similar patterns of build-up of protection after the first dose and waning of protection after the second dose. The relative differences between those vaccinated with mRNA-1273 versus BNT162b2 were stable over time after vaccination. For both vaccines, most breakthrough infections after the first dose occurred in the first two weeks after vaccination. Also, for both vaccines, most breakthrough infections after the second dose occurred 90 days or more after the second dose, consistent with similar waning of vaccine immunity for both vaccines. These results may suggest that the nature of immunity that builds after vaccination and wanes over time is similar for both vaccines, but that the differences in the level of protection could be due to differences in the magnitude (not mechanistic type) of initial activation of immune response following the first and second doses. These findings may be explained by the larger dose of the mRNA-1273 vaccine versus that of the BNT162b2 vaccine.^1,2^ Evidence suggests that the mRNA-1273 vaccine induces higher neutralizing antibody titers than the BNT162b2 vaccine.^4^ The interval between first and second doses is one week longer for the mRNA-1273 vaccine, and evidence suggests greater protection with longer interval between doses.^33^

This study has the following limitations. Vaccinated cohorts in Qatar predominantly included young and working-age adults; thus, these findings may not generalize to children or older persons. As an observational study, the vaccinated cohorts were neither blinded nor randomized, so the potential for unmeasured or uncontrolled confounding cannot be excluded. While matching was done for age, sex, nationality, and calendar week of the first (or second) vaccine dose, it was not possible for other factors, such as comorbidities or occupation, as these data were not available to study investigators.

However, matching was done to control for major factors known to affect risk of exposure to the infection in Qatar,^14,17,26-28^ as well as variant exposure over the course of follow-up.^6,8,9,19,23-25^ Matching by age may have (partially) reduced the potential for bias due to co-morbidities. The number of individuals with severe chronic conditions is also small in Qatar’s young population.^14,34^ Matching by nationality may have (partially) controlled the differences in occupational risk, in consideration of the association between nationality and occupation in Qatar.^14,17,26-28^ Both vaccines were broadly distributed across Qatar’s neighborhoods/areas and population social substrata. Individuals were vaccinated by the vaccine that was available at the time of the vaccination appointment. Cohorts were matched by calendar week of the first (or second) dose; thus, vaccination eligibility criteria were the same for both vaccines at the time of vaccination. PCR testing rates were similar for both the mRNA-1273- and BNT162b2-vaccinated cohorts. Importantly, there was no difference in infection incidence in the first two weeks after the first dose, as expected, considering evidence of negligible vaccine protection in these two weeks.^1,2,6-8,19^ This supports adequate control of potential bias arising from differences in the risk of exposure to the infection between these vaccinated cohorts.

In conclusion, mRNA-1273 vaccination is associated with significantly lower SARS-CoV-2 breakthrough infection and severe COVID-19 than BNT162b2 vaccination. Despite these differences, both vaccines elicited robust protection against COVID-19 hospitalization and death. Both vaccines also demonstrated remarkably similar patterns of build-up of protection after the first dose and waning of protection after the second dose. The nature of vaccine immunity that builds after vaccination and wanes over time appears similar for both vaccines. Differences in the level of protection perhaps arise from the differences in the size of the vaccine dose.

## Data Availability

The dataset of this study is a property of the Qatar Ministry of Public Health that was provided to the researchers through a restricted-access agreement that prevents sharing the dataset with a third party or publicly. Future access to this dataset can be considered through a direct application for data access to Her Excellency the Minister of Public Health (https://www.moph.gov.qa/english/Pages/default.aspx). Aggregate data are available within the manuscript and its Supplementary information.

## Acknowledgements

We acknowledge the many dedicated individuals at Hamad Medical Corporation, the Ministry of Public Health, the Primary Health Care Corporation, Qatar Biobank, Sidra Medicine, and Weill Cornell Medicine-Qatar for their diligent efforts and contributions to make this study possible. The authors are grateful for institutional salary support from the Biomedical Research Program and the Biostatistics, Epidemiology, and Biomathematics Research Core, both at Weill Cornell Medicine-Qatar, as well as for institutional salary support provided by the Ministry of Public Health and Hamad Medical Corporation. The authors are also grateful for the Qatar Genome Programme for institutional support for the reagents needed for the viral genome sequencing. The funders of the study had no role in study design, data collection, data analysis, data interpretation, or writing of the article. Statements made herein are solely the responsibility of the authors.

## Author contributions

LJA conceived and co-designed the study, led the statistical analyses, and co-wrote the first draft of the article. HC co-designed the study, performed the statistical analyses, and co-wrote the first draft of the article. PT and MRH conducted the multiplex, RT-qPCR variant screening and viral genome sequencing. HY, FMB, and HAK conducted viral genome sequencing. All authors contributed to data collection and acquisition, database development, discussion and interpretation of the results, and to the writing of the manuscript. All authors have read and approved the final manuscript.

## Competing interests

Dr. Butt has received institutional grant funding from Gilead Sciences unrelated to the work presented in this paper. Otherwise, we declare no competing interests.

## Supplementary Appendix

### Section 1. COVID-19 severity, criticality, and fatality classification

Severe Coronavirus Disease 2019 (COVID-19) disease was defined per the World health Organization (WHO) classification as a severe acute respiratory syndrome coronavirus 2 (SARS-CoV-2) infected person with “oxygen saturation of <90% on room air, and/or respiratory rate of >30 breaths/minute in adults and children >5 years old (or ≥60 breaths/minute in children <2 months old or ≥50 breaths/minute in children 2-11 months old or ≥40 breaths/minute in children 1–5 years old), and/or signs of severe respiratory distress (accessory muscle use and inability to complete full sentences, and, in children, very severe chest wall indrawing, grunting, central cyanosis, or presence of any other general danger signs)”.^1^ Detailed WHO criteria for classifying SARS-CoV-2 infection severity can be found in the WHO technical report.^1^

Critical COVID-19 disease was defined per WHO classification as a SARS-CoV-2 infected person with “acute respiratory distress syndrome, sepsis, septic shock, or other conditions that would normally require the provision of life sustaining therapies such as mechanical ventilation (invasive or non-invasive) or vasopressor therapy”.^1^ Detailed WHO criteria for classifying SARS-CoV-2 infection criticality can be found in the WHO technical report.^1^

COVID-19 death was defined per WHO classification as “a death resulting from a clinically compatible illness, in a probable or confirmed COVID-19 case, unless there is a clear alternative cause of death that cannot be related to COVID-19 disease (e.g. trauma). There should be no period of complete recovery from COVID-19 between illness and death. A death due to COVID-19 may not be attributed to another disease (e.g. cancer) and should be counted independently of preexisting conditions that are suspected of triggering a severe course of COVID-19”. Detailed WHO criteria for classifying COVID-19 death can be found in the WHO technical report.^2^

### Section 2. Laboratory methods

Nasopharyngeal and/or oropharyngeal swabs were collected for PCR testing and placed in Universal Transport Medium (UTM). Aliquots of UTM were: 1) extracted on a QIAsymphony platform (QIAGEN, USA), KingFisher Flex (Thermo Fisher Scientific, USA), MGISP-960 (MGI, China), or ExiPrep 96 Lite (Bioneer, South Korea) followed by testing with real-time reverse-transcription PCR (RT-qPCR) using TaqPath(tm) COVID-19 Combo Kits (Thermo Fisher Scientific, USA) on an ABI 7500 FAST (Thermo Fisher Scientific, USA); 2) tested directly on the Cepheid GeneXpert system using the Xpert Xpress SARS-CoV-2 (Cepheid, USA); or 3) loaded directly into a Roche cobas® 6800 system and assayed with the cobas® SARS-CoV-2 Test (Roche, Switzerland). The first assay targets the viral S, N, and ORF1ab gene regions. The second targets the viral N and E-gene regions, and the third targets the ORF1ab and E-gene regions.

All PCR testing was conducted at the Hamad Medical Corporation Central Laboratory or Sidra Medicine Laboratory, following standardized protocols.

### Section 3. Classification of infections by variant type

Surveillance for SARS-CoV-2 variants in Qatar is based on viral genome sequencing and multiplex, real-time reverse-transcription PCR (RT-qPCR) variant screening^3^ of random positive clinical samples,^4-9^ and complemented by deep sequencing of wastewater samples.^6,10^ The latter is used to compare the distribution of variants in wastewater with that in clinical samples collected from SARS-CoV-2 patients.

Between March 23, 2021 and October 9, 2021, RT-qPCR genotyping identified 6,418 (31.9%) Beta (B.1.351)-like cases, 4,134 (20.6%) Alpha (B.1.1.7)-like cases, 9,499 (47.2%) “other” variant cases, and 61 (0.3%) B.1.375-like or B.1.258-like cases in 20,112 randomly collected SARS-CoV-2-positive specimens.^6,8^

The accuracy of the RT-qPCR genotyping was verified against either Sanger sequencing of the receptor-binding domain (RBD) of SARS-CoV-2 surface glycoprotein (S) gene, or by viral whole-genome sequencing on a Nanopore GridION sequencing device. From 236 random samples (27 Alpha-like, 186 Beta-like, and 23 “other” variants), PCR genotyping results for Alpha-like, Beta-like, and ‘other’ variants were in 88.8% (23 out of 27), 99.5% (185 out of 186), and 100% (23 out of 23) agreement with the SARS-CoV-2 lineages assigned by sequencing. Within the “other” variant category, Sanger sequencing and/or Illumina sequencing of the RBD of SARS-CoV-2 spike gene on 728 random samples confirmed that 701 (96.3%) were Delta cases and 17 (2.3%) were other variant cases, with 10 (1.4%) samples failing lineage assignment.^6,8^ Accordingly, a Delta case was proxied as any “other” case identified through the RT-qPCR based variant screening.

All the variant RT-qPCR screening was conducted at the Sidra Medicine Laboratory following standardized protocols.

---

**Table S1.** STROBE checklist for cohort studies.

|  | Item No | Recommendation | Main Text page no |
| --- | --- | --- | --- |
| Title and abstract | 1 | (a) Indicate the study’s design with a commonly used term in the title or the abstract | 2 |
|  |  | (b) Provide in the abstract an informative and balanced summary of what was done and what was found | 2-3 |
| Introduction |  |  |  |
| Background/rationale | 2 | Explain the scientific background and rationale for the investigation being reported | 4 |
| Objectives | 3 | State specific objectives, including any prespecified hypotheses | 4 |
| Methods |  |  |  |
| Study design | 4 | Present key elements of study design early in the paper | 5-6 |
| Setting | 5 | Describe the setting, locations, and relevant dates, including periods of recruitment, exposure, follow-up, and data collection | 5-6 & Sections 1-3 |
| Participants | 6 | (a) Give the eligibility criteria, and the sources and methods of selection of participants. Describe methods of follow-up<br>(b) For matched studies, give matching criteria and number of exposed and unexposed | 5-7, Figure 1 & Sections 1-3 |
| Variables | 7 | Clearly define all outcomes, exposures, predictors, potential confounders, and effect modifiers. Give diagnostic criteria, if applicable | 5-7, Figure 1, Tables 1-3, & Sections 1-3 |
| Data sources/measurement | 8* | For each variable of interest, give sources of data and details of methods of assessment (measurement). Describe comparability of assessment methods if there is more than one group | 5-7, Sections 1-3, & Tables 1-3 |
| Bias | 9 | Describe any efforts to address potential sources of bias | 5 & 7 |
| Study size | 10 | Explain how the study size was arrived at | Figure 1 |
| Quantitative variables | 11 | Explain how quantitative variables were handled in the analyses. If applicable, describe which groupings were chosen and why | 7 & Tables 1-3 |
| Statistical methods | 12 | (a) Describe all statistical methods, including those used to control for confounding | 7 |
|  |  | (b) Describe any methods used to examine subgroups and interactions | 7 |
|  |  | (c) Explain how missing data were addressed | NA, see p. 5 |
|  |  | (d) If applicable, explain how loss to follow-up was addressed | NA |
|  |  | (e) Describe any sensitivity analyses | NA |
| Results |  |  |  |
| Participants | 13* | (a) Report numbers of individuals at each stage of study—eg numbers potentially eligible, examined for eligibility, confirmed eligible, included in the study, completing follow-up, and analysed<br>(b) Give reasons for non-participation at each stage<br>(c) Consider use of a flow diagram | Figure 1, Tables 1-2 |
| Descriptive data | 14 | (a) Give characteristics of study participants (eg demographic, clinical, social) and information on exposures and potential confounders | 7-10, & Tables 1-3 |
|  |  | (b) Indicate number of participants with missing data for each variable of interest | NA, see p. 5 |
|  |  | (c) Summarise follow-up time (eg, average and total amount) | Tables 1-2, Figure 2 |
| Outcome data | 15 | Report numbers of outcome events or summary measures over time | 9-10, Figure 1, Table 3-4 |
| Main results | 16 | (a) Give unadjusted estimates and, if applicable, confounder-adjusted estimates and their precision (eg, 95% confidence interval). Make clear which confounders were adjusted for and why they were included | 9-12, Figure 2, & Table 4 |
|  |  | (b) Report category boundaries when continuous variables were categorized | Tables 1-3 |
|  |  | (c) If relevant, consider translating estimates of relative risk into absolute risk for a meaningful time period | NA |
| Other analyses | 17 | Report other analyses done—eg analyses of subgroups and interactions, and sensitivity analyses | 10-11 & Table 4 |
| Discussion |  |  |  |
| Key results | 18 | Summarise key results with reference to study objectives | 12-13 |
| Limitations | 19 | Discuss limitations of the study, taking into account sources of potential bias or imprecision. Discuss both direction and magnitude of any potential bias | 13-14 |
| Interpretation | 20 | Give a cautious overall interpretation of results considering objectives, limitations, multiplicity of analyses, results from similar studies, and other relevant evidence | 14 |
| Generalisability | 21 | Discuss the generalisability (external validity) of the study results | 13 |
| Other information |  |  |  |
| Funding | 22 | Give the source of funding and the role of the funders for the present study and, if applicable, for the original study on which the present article is based | Acknowledgements |

Abbreviations: NA, not applicable.

## Notes

### Competing Interest Statement

The authors have declared no competing interest.

## References

1. Baden LR, El Sahly HM, Essink B, et al. Efficacy and Safety of the mRNA-1273 SARS-CoV-2 Vaccine. N Engl J Med 2021;384:403–16.

2. Polack FP, Thomas SJ, Kitchin N, et al. Safety and Efficacy of the BNT162b2 mRNA Covid-19 Vaccine. N Engl J Med 2020.

3. Vaccine effectievness studies. VIEW-hub. International Vaccine Access Center. https://view-hub.org/covid-19/effectiveness-studies/. Accessed on October 26, 2021.

4. Khoury DS, Cromer D, Reynaldi A, et al. Neutralizing antibody levels are highly predictive of immune protection from symptomatic SARS-CoV-2 infection. Nat Med 2021;27:1205–11.

5. Nordström P, Ballin M, Nordström A. Effectiveness of Covid-19 Vaccination Against Risk of Symptomatic Infection, Hospitalization, and Death Up to 9 Months: A Swedish Total-Population Cohort Study. Available at SSRN: https://ssrn.com/abstract=3949410. xpreprint 2021.

6. Abu-Raddad LJ, Chemaitelly H, Butt AA, National Study Group for Covid-19 Vaccination. Effectiveness of the BNT162b2 Covid-19 Vaccine against the B.1.1.7 and B.1.351 Variants. N Engl J Med 2021.

7. Abu-Raddad LJ, Chemaitelly H, Yassine HM, et al. Pfizer-BioNTech mRNA BNT162b2 Covid-19 vaccine protection against variants of concern after one versus two doses. J Travel Med 2021.

8. Chemaitelly H, Yassine HM, Benslimane FM, et al. mRNA-1273 COVID-19 vaccine effectiveness against the B.1.1.7 and B.1.351 variants and severe COVID-19 disease in Qatar. Nat Med 2021;27:1614–21.

9. Tang P, Hasan MR, Chemaitelly H, et al. BNT162b2 and mRNA-1273 COVID-19 vaccine effectiveness against the Delta (B.1.617.2) variant in Qatar. In press at Nature Medicine. medRxiv 2021:2021.08.11.21261885.

10. Abu-Raddad LJ, Chemaitelly H, Ayoub HH, et al. Protection afforded by the BNT162b2 and mRNA-1273 COVID-19 vaccines in fully vaccinated cohorts with and without prior infection. In press at JAMA. medRxiv 2021:2021.07.25.21261093.

11. Abu-Raddad LJ, Chemaitelly H, Ayoub HH, et al. Effect of vaccination and of prior infection on infectiousness of vaccine breakthrough infections and reinfections. medRxiv 2021:2021.07.28.21261086.

12. World Health Organization. COVID-19 clinical management: living guidance. Available from: https://www.who.int/publications/i/item/WHO-2019-nCoV-clinical-2021-1. Accessed on: May 15 2021. 2021.

13. World Health Organization. International guidelines for certification and classification (coding) of COVID-19 as cause of death. Available from: https://www.who.int/classifications/icd/Guidelines_Cause_of_Death_COVID-19-20200420-EN.pdf?ua=1. Document Number: WHO/HQ/DDI/DNA/CAT. Accessed on May 31, 2021. 2021.

14. Abu-Raddad LJ, Chemaitelly H, Ayoub HH, et al. Characterizing the Qatar advanced-phase SARS-CoV-2 epidemic. Sci Rep 2021;11:6233.

15. Abu-Raddad LJ, Chemaitelly H, Coyle P, et al. SARS-CoV-2 antibody-positivity protects against reinfection for at least seven months with 95% efficacy. EClinicalMedicine 2021;35:100861.

16. Abu-Raddad LJ, Chemaitelly H, Malek JA, et al. Assessment of the risk of SARS-CoV-2 reinfection in an intense re-exposure setting. Clin Infect Dis 2020.

17. Ayoub HH, Chemaitelly H, Seedat S, et al. Mathematical modeling of the SARS-CoV-2 epidemic in Qatar and its impact on the national response to COVID-19. J Glob Health 2021;11:05005.

18. Bertollini R, Chemaitelly H, Yassine HM, Al-Thani MH, Al-Khal A, Abu-Raddad LJ. Associations of Vaccination and of Prior Infection With Positive PCR Test Results for SARS-CoV-2 in Airline Passengers Arriving in Qatar. JAMA 2021.

19. Chemaitelly H, Tang P, Hasan MR, et al. Waning of BNT162b2 Vaccine Protection against SARS-CoV-2 Infection in Qatar. N Engl J Med 2021.

20. Planning and Statistics Authority-State of Qatar. Qatar Monthly Statistics. Available from: https://www.psa.gov.qa/en/pages/default.aspx. Accessed on: May 26, 2020. 2020.

21. Ministry of Public Health. National Covid-19 Testing and Vaccination Program Data. https://covid19.moph.gov.qa/EN/Pages/Vaccination-Program-Data.aspx. Access date October 26, 2021. 2021.

22. World Health Organization. Tracking SARS-CoV-2 variants. Available from: https://www.who.int/en/activities/tracking-SARS-CoV-2-variants/. Accessed on: June 5, 2021. 2021.

23. Qatar viral genome sequencing data. Data on randomly collected samples. https://www.gisaid.org/phylodynamics/global/nextstrain/. 2021. at https://www.gisaid.org/phylodynamics/global/nextstrain/.)

24. Benslimane FM, Al Khatib HA, Al-Jamal O, et al. One year of SARS-CoV-2: Genomic characterization of COVID-19 outbreak in Qatar. medRxiv 2021:2021.05.19.21257433.

25. Hasan MR, Kalikiri MKR, Mirza F, et al. Real-Time SARS-CoV-2 Genotyping by High-Throughput Multiplex PCR Reveals the Epidemiology of the Variants of Concern in Qatar. Int J Infect Dis 2021.

26. Coyle PV, Chemaitelly H, Kacem M, et al. SARS-CoV-2 seroprevalence in the urban population of Qatar: An analysis of antibody testing on a sample of 112,941 individuals. iScience 2021:102646.

27. Al-Thani MH, Farag E, Bertollini R, et al. SARS-CoV-2 infection is at herd immunity in the majority segment of the population of Qatar. Open Forum Infectious Diseases 2021.

28. Jeremijenko A, Chemaitelly H, Ayoub HH, et al. Herd Immunity against Severe Acute Respiratory Syndrome Coronavirus 2 Infection in 10 Communities, Qatar. Emerg Infect Dis 2021;27:1343–52.

29. Multiplexed RT-qPCR to screen for SARS-COV-2 B.1.1.7, B.1.351, and P.1 variants of concern V.3. dx.doi.org/10.17504/protocols.io.br9vm966. 2021. (Accessed June 6, 2021, at https://www.protocols.io/view/multiplexed-rt-qpcr-to-screen-for-sars-cov-2-b-1-1-br9vm966.)

30. Austin PC. Using the Standardized Difference to Compare the Prevalence of a Binary Variable Between Two Groups in Observational Research. Communications in Statistics - Simulation and Computation 2009;38:1228–34.

31. Kaplan EL, Meier P. Nonparametric estimation from incomplete observations. J Amer Statist Assoc 1958;53:457–81.

32. StataCorp. Stata Statistical Software: Release 17. College Station, TX: StataCorp LLC. 2021.

33. Voysey M, Costa Clemens SA, Madhi SA, et al. Single-dose administration and the influence of the timing of the booster dose on immunogenicity and efficacy of ChAdOx1 nCoV-19 (AZD1222) vaccine: a pooled analysis of four randomised trials. Lancet 2021;397:881–91.

34. Seedat S, Chemaitelly H, Ayoub HH, et al. SARS-CoV-2 infection hospitalization, severity, criticality, and fatality rates in Qatar. Sci Rep 2021;11:18182.

## References

1. World Health Organization. COVID-19 clinical management: living guidance. Available from: https://www.who.int/publications/i/item/WHO-2019-nCoV-clinical-2021-1. Accessed on: May 15 2021. 2021.

2. World Health Organization. International guidelines for certification and classification (coding) of COVID-19 as cause of death. Available from: https://www.who.int/classifications/icd/Guidelines_Cause_of_Death_COVID-19-20200420-EN.pdf?ua=1. Document Number: WHO/HQ/DDI/DNA/CAT. Accessed on May 31, 2021. 2021.

3. Multiplexed RT-qPCR to screen for SARS-COV-2 B.1.1.7, B.1.351, and P.1 variants of concern V.3. dx.doi.org/10.17504/protocols.io.br9vm966. 2021. (Accessed June 6, 2021, at https://www.protocols.io/view/multiplexed-rt-qpcr-to-screen-for-sars-cov-2-b-1-1-br9vm966.)

4. Abu-Raddad LJ, Chemaitelly H, Butt AA, National Study Group for Covid-19 Vaccination. Effectiveness of the BNT162b2 Covid-19 Vaccine against the B.1.1.7 and B.1.351 Variants. N Engl J Med 2021.

5. Chemaitelly H, Yassine HM, Benslimane FM, et al. mRNA-1273 COVID-19 vaccine effectiveness against the B.1.1.7 and B.1.351 variants and severe COVID-19 disease in Qatar. Nat Med 2021.

6. Qatar viral genome sequencing data. Data on randomly collected samples. https://www.gisaid.org/phylodynamics/global/nextstrain/. 2021. at https://www.gisaid.org/phylodynamics/global/nextstrain/.)

7. Benslimane FM, Al Khatib HA, Al-Jamal O, et al. One year of SARS-CoV-2: Genomic characterization of COVID-19 outbreak in Qatar. medRxiv 2021:2021.05.19.21257433.

8. Hasan MR, Kalikiri MKR, Mirza F, et al. Real-Time SARS-CoV-2 Genotyping by High-Throughput Multiplex PCR Reveals the Epidemiology of the Variants of Concern in Qatar. Int J Infect Dis 2021.

9. Chemaitelly H, Tang P, Hasan MR, et al. Waning of BNT162b2 Vaccine Protection against SARS-CoV-2 Infection in Qatar. N Engl J Med 2021.

10. Saththasivam J, El-Malah SS, Gomez TA, et al. COVID-19 (SARS-CoV-2) outbreak monitoring using wastewater-based epidemiology in Qatar. Sci Total Environ 2021;774:145608.

